# Increase but pronounced regional disparities in gamma-hydroxybutyrate (GHB) prescriptions among Medicaid and Medicare patients

**DOI:** 10.1101/2024.02.20.24303095

**Authors:** Josephine R. Barnhart, Sondra A. Vujovich, Brian J. Piper

## Abstract

**Background:** Gamma-hydroxybutyrate (GHB) is a Schedule III drug in the US approved for treatment of cataplexy associated with narcolepsy. Narcolepsy with cateplaxy is a rare disorder with an annual incidence of less than one per one-hundred thousand and GHB is a third-line treatment. The purpose of this study was to describe the temporal pattern of GHB distribution and cost nationally and between states for Medicaid and Medicare patients.

**Methods:** GHB prescriptions were extracted from the State Utilization Data Tool from Medicaid.gov and the Medicare part D Prescribers by Provider and Drug Dataset from CMS.gov. GHB prescriptions were examined by state when corrected for population. States outside a 95% confidence interval were considered statistically significant. GHB cost analyses were performed between 2017-2021. GHB prescribers were identified for Medicare in 2019.

**Results:** There was a steady increase in prescriptions (+88.5%) from 2019 to 2021 and spending (+39.6%) from 2017 to 2020 for Medicaid. Specialists other than somnologists, were found to prescribe the highest number of GHB prescriptions to Medicare Part D enrollees. In 2019, two states (Hawaii and North Dakota) did not prescribe GHB to Medicare patients versus twenty states for Medicaid patients. Maryland’s prescribing to Medicare patients was significantly elevated (269.2/100K).

**Conclusion:** GHB prescribing has increased to Medicaid and Medicare patients. Further research is necessary to understand how the COVID-19 pandemic and off-label prescribing (e.g. for excessive daytime sleepiness) may have affected these findings including the origins of the pronounced state level disparities.

## Introduction

Gamma-hydroxybutyrate (GHB) is the sodium salt of gamma-hydroxybutyric acid, an inhibitory neurotransmitter (1). GHB (Xyrem, also known as sodium oxybate) was FDA approved in July, 2002 in adults for daytime sleepiness and muscle weakness with narcolepsy, denoting a sudden loss of muscle tone while awake (2) and for children in October 2018. An oral solution formulation containing sodium, potassium, magnesium, and calcium oxybates was also approved for cataplexy and/or excessive daytime sleepiness in narcolepsy, aged ≥ 7 in July, 2020 and for idiopathic hypersomnia (IH) in adults in August of 2021. A population-based study in Olmsted County, Minnesota estimated that the prevalence of narcolepsy with cataplexy at one in 2,793 people (i.e. 33.6 per 100,000) and the incidence was 0.74 per 100,000 (3). Narcolepsy treatment guidelines list modafinil/armodafinil, pitolisant, and solriamfetol as first-line; methylphenidate and amphetamines as second-line, and the addition of oxybates as third-line (4). The prevalence of IH is unknown but it is estimated at 2-5 cases per 100,000 (5). Modafinil is the first-line therapy for IH and oxybates are second-line (5). In addition to GHB’s use for daytime sleepiness (6), it is prescribed off-label for alcohol use disorder treatment (7). Jazz pharmaceuticals paid over twenty million in fines for inappropriate marketing for insomnia, depression, and fibromyalgia (8). Potential risk of misuse of this Schedule III substance led to creation of the Xyrem US Risk Evaluation and Mitigation Program which incorporates strict guidelines to monitor, prescriber and patient enrollment, education, documentation, drug usage and dispensing, and pharmacists’ certification (9). There was a 29.1% increase among an international self-selected sample in recreational GHB/gamma butyrolactone (i.e. the precursor of GHB) drug consumption during the COVID-19 pandemic (10). Sodium oxybate was ranked the second drug for the most number of payments made by pharmaceutical companies to sleep medicine (11). There have also been concerns about newer GHB formulations costing over one-hundred thousand per patient per year (8) for pharmacotherapies that are not disease modifying (4).

This investigation examined the change in usage of GHB among US Medicaid and Medicare patients. State level differences in prescribing were examined as has previously been completed for other psychoactive substances (12, 13, 14, 15). Inhomogeneity in prescribing across states may be due to a variety of factors including differences in disease prevalence, preferred drug lists, and program enrollment criteria. If use of this non-first line pharmacotherapy was for FDA approved indications, then we anticipated that prescribing would be less than the prevalence of these conditions.

## Methods

### Databases

Data was extracted from the Medicaid State Utilization and the Medicare Part D Prescribers and Drug claims databases for generic and brand name formulations (Xyrem, Xywav, Lumryz). Medicaid.gov provided breakdowns of the amount and cost of drugs being prescribed to Medicaid enrollees (16). GHB claims and spending were also extracted for each state (16). Similarly, Data.CMS.gov provided the claims and cost of GHB prescribed to the Medicare enrollees (17). The Medicare prescription database also provided what type of medical provider was prescribing GHB (17). Data from the District of Columbia was incorporated into the Maryland data to keep both databases consistent. Procedures were approved as exempt by the IRB of Geisinger.

### Procedures

Medicaid.gov provided GHB claims for 2017-2021. Data.CMS.gov provided the following variables: number of GHB prescriptions per year, number of GHB prescriptions per state, and type of GHB prescription prescriber. Prescriptions for Schedule III substances cannot be for longer than six-months. Medicare prescriber data was obtained for 2019 (i.e. the most recent complete year available when data was extracted). The number of prescriptions were corrected based on the number of Medicaid/CHIP or Medicare enrollees. Extraction of Medicaid was on August, 2023 and Medicare data extraction was July, 2023. Data were suppressed from states who reported less than 11 GHB prescriptions for a National Drug Code per quarter as per the CMS confidentiality policy.

### Data Analysis

Graphs and figures were constructed from Microsoftl71 Excel version 2022 for Windows, GraphPad Prism, and JMP statistical software. States with prescribing rates outside of a 95% confidence interval (mean ± 1.96* SD) were considered statistically significant.^7-10^ Percent differences in cost of GHB prescriptions from 2017-2021were also reported. A Pearson correlation between state level prescribing in Medicaid and Medicare was completed.

## Results

Medicaid prescribing nationally increased by 89.2% from 2019 (9.37/100K enrollees) to 2021 (11.0 / 100K). Similarly, spending increased 107.6% from 2017 ($85,316,479.90 million) to 2021 ($177,087,312.21 million). Figure 1 shows a 10.5 fold difference between the highest (Indiana = 50.2 / 100K) and lowest non-zero (Rhode Island = 4.8 / 100K) states in 2019. Indiana (50.2), Connecticut (35.0), and Ohio (34.6) were significantly (p < .05) elevated relative to the national average (10.3). Two-fifths of states including Maryland had no prescribing. There was a 19.2 fold difference between the highest (Virginia = 39.4 / 100K) and lowest non-zero (South Carolina = 2.05 / 100K) states in 2021. Virginia was the sole state that was significantly (p < .05) elevated relative to the national average (11.0). Similarly to 2019, two-fifths of states had no prescribing (Figure 2).

**Figure 1.**
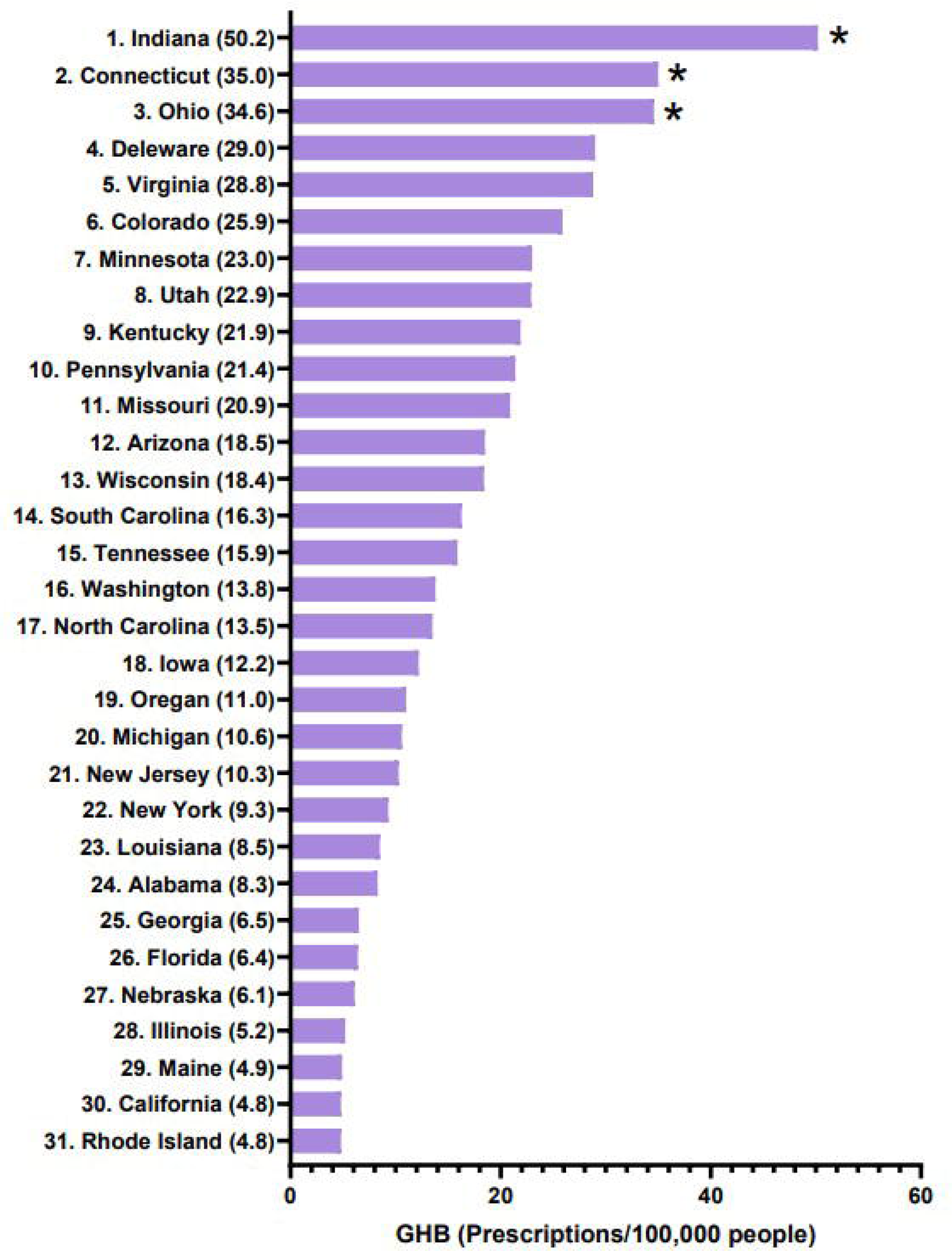
GHB prescriptions per one-hundred thousand Medicaid enrollees in 2019. Nineteen states (AK, AR, HI, ID, KS, MA, MD, MS, MT, ND, NH, NM, NV, OK, SD, TX, VT, WV, WY) are not displayed due to no (<11 per National Drug Code) prescriptions. * p < .05 vs the average (10.3).

**Figure 2:**
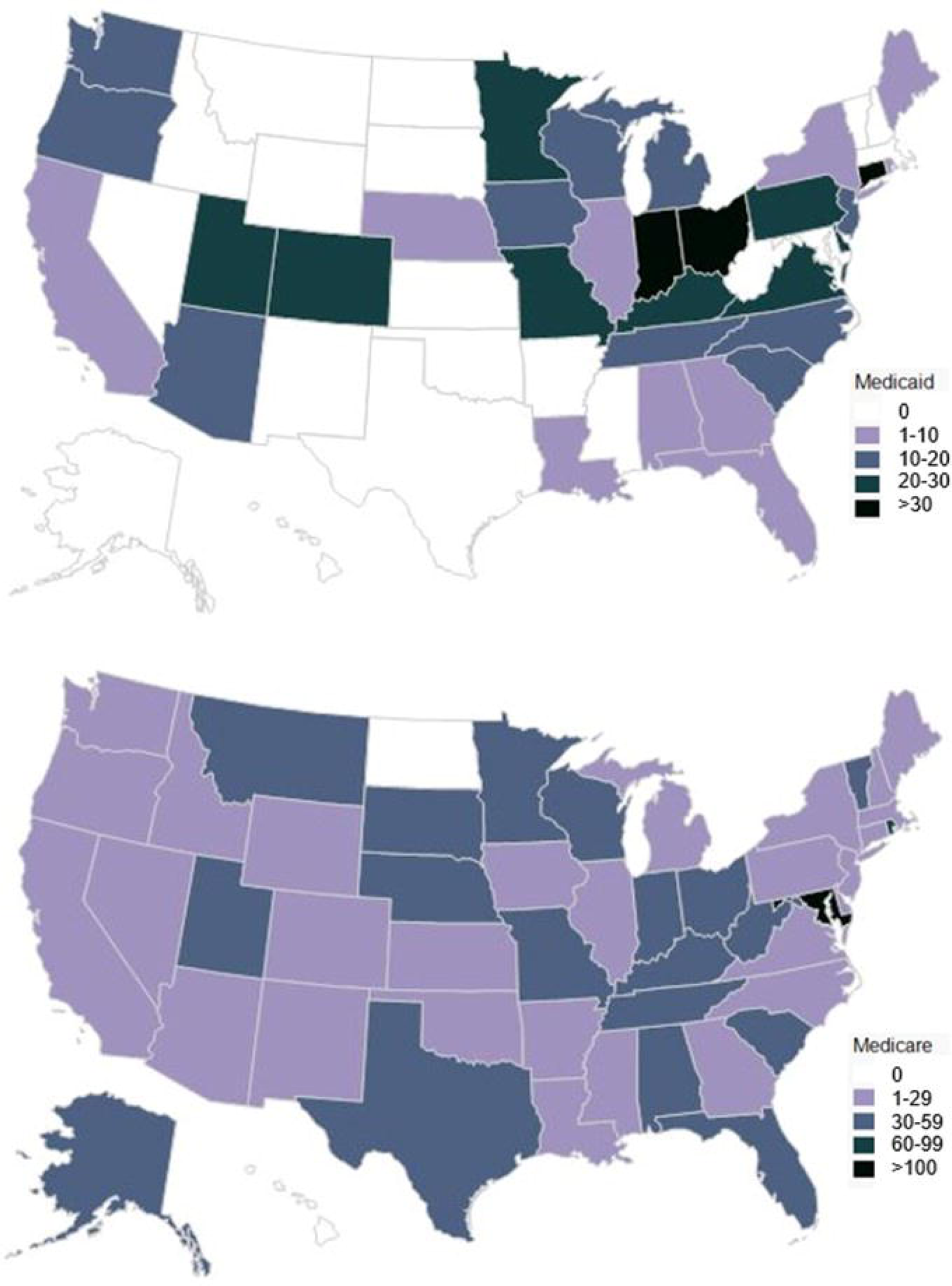
GHB prescriptions per one-hundred thousand Medicaid enrollees in 2021. Sixteen states (AK, AR, DE,HI, MA, MD, MS, MT, ND, NH, NM, SD, TX, VT, WV, WY) are not listed due to no (<11 per National Drug Code) prescriptions. * p < .05 vs the average (16.2).

Medicare prescribing in 2019 nationally (16,498 prescriptions) was 2.1 fold higher than Medicaid (7,929 prescriptions). Prescribing in Maryland (269.2 / 100K) was significantly elevated and 4.5 fold higher than the next highest state (Rhode Island = 59.3) (Figure 3). Contrary to Medicaid, Medicare saw a decrease (-19.4%) in total number of GHB being prescribed from 2019 to 2021 (Figure 4). Further analyses were completed on who was prescribing in 2019. In Maryland, pulmonary made up 51.3% of prescribers followed by neurology (25.3%), nurse practitioners (19.2%), and sleep medicine (4.2%). On a national level, Wyoming was the lowest prescribing state (neurologists being the sole prescribers at .08% of the national total of prescriptions) versus Florida being the highest prescribing state (9.2%).

**Figure 3.**
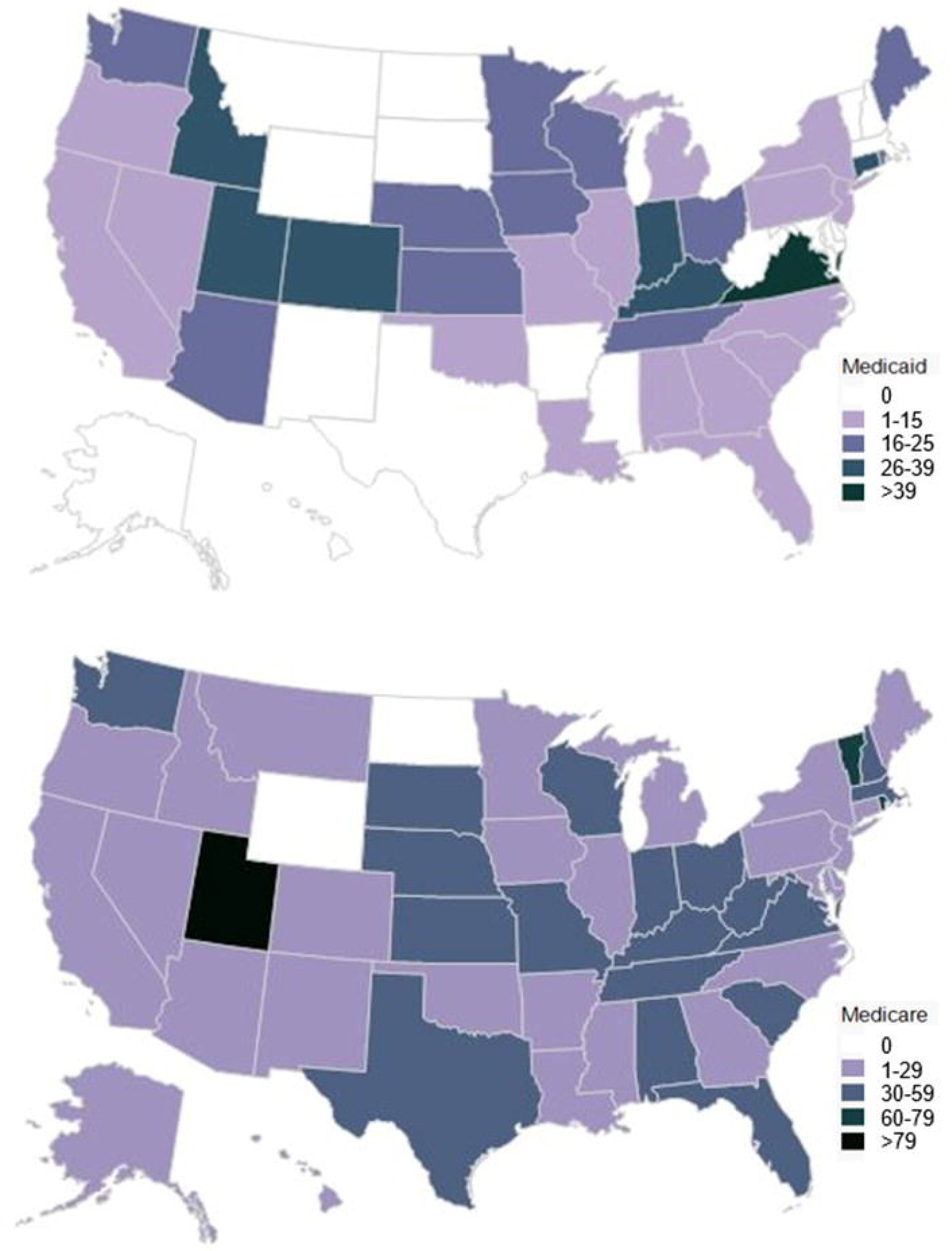
GHB prescriptions per one-hundred thousand Medicare Part D enrollees in 2019. *p < .05 vs the state average (31.6).

**Figure 4.**
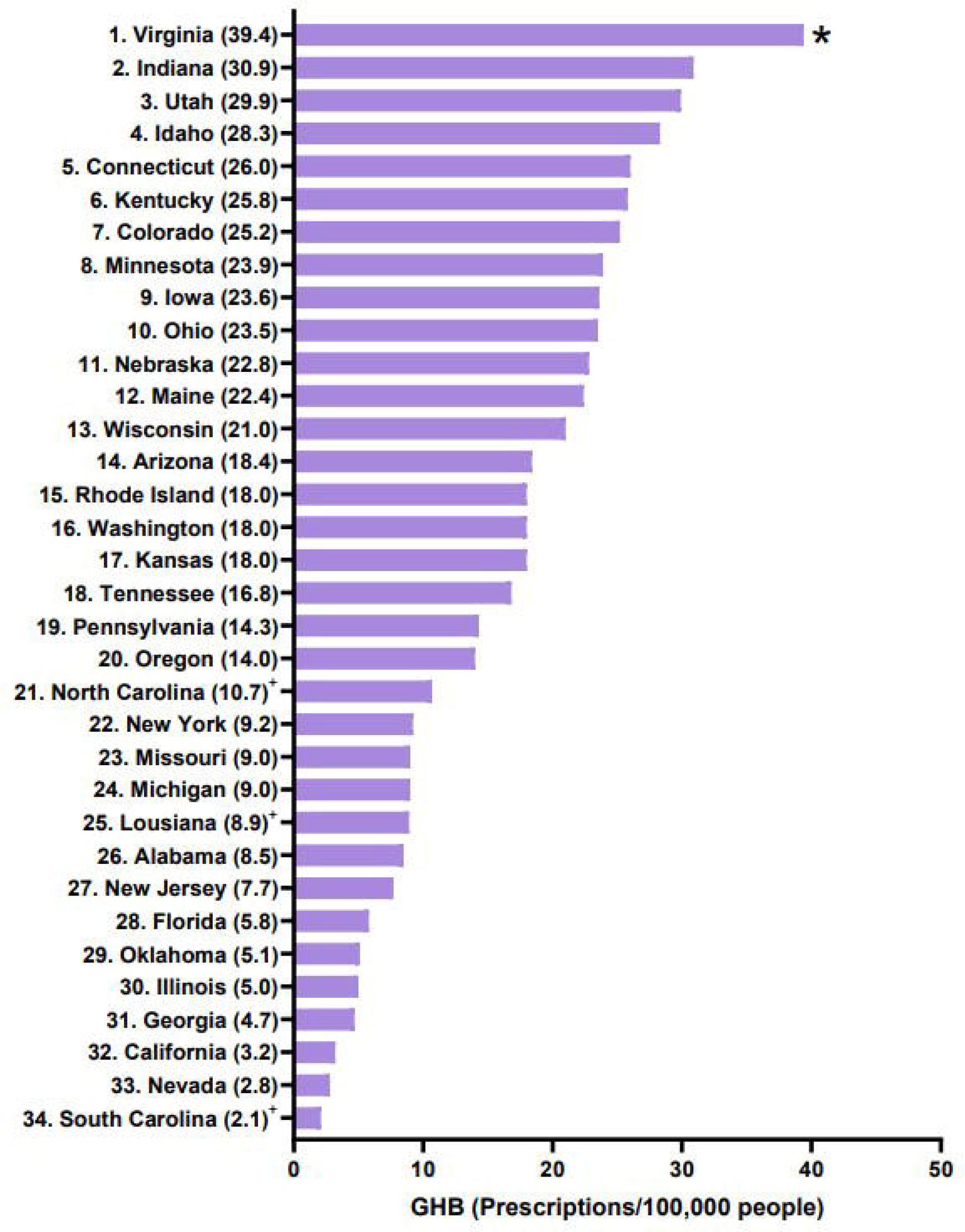
GHB prescriptions per one-hundred Medicare Part D enrollees in 2019. *p < .05 vs the state average (30.4).

## Discussion

There are three findings of this novel report. First, Medicaid saw an increase in GHB prescribing (+89.2%) from 2019 to 2021 and spending (+39.6%) increased to Medicaid patients from 2017 to 2021. Additionally, an overall decrease of -19.4% in Medicare GHB prescriptions were seen from 2019-2021. This study also identified pronounced state level differences among both Medicaid and Medicare patients. Medicaid serves primarily a resource limited population (age < 65) and Medicare serves primarily an older (age ≥ 65) population. Rather than being used solely as a narcolepsy drug, GHB has been found to be used in cardiology as a long-term sedative without depression of cardio-circulatory or respiratory measures, kidney and other functions (18). The mean age for bypass patients undergoing heart surgery was 68.5 years (19), a demographic that correlates to Medicare Part D enrollees’ outpatient prescription drug benefits program (20).

An important implication to consider for this study and future research is the issue of drug misuse. Despite this study covering GHB prescriptions that were prescribed by a health care provider, there is still concern about the prescriptions being inappropriately used by patients, as well as being inappropriately prescribed to GHB drug misusers for non-medical purposes. Despite majority of drug consumption showing a decrease in usage, GHB users experienced the highest increase in drug usage (21). These observations were due to GHB users reporting cost and obtainability of GHB to be one of the least expensive and easiest drugs to obtain (21).

There are also several limitations to consider. First, the main unit of analysis was prescriptions. A single patient could receive two or more prescriptions in a single year. These findings are applicable for Medicaid and Medicare patients which account for over one-third of the US population but further study among those with private insurance should be completed. State-level differences in prescribing are as identified when data was extracted but the CMS databases may be updated by states in the future. As narcolepsy is an uncommon disorder (19-56/100,000 people), additional study with electronic medical records may be necessary to determine what percent of prescriptions were for off-label indications (22).

In conclusion, this study found a significant state-level differences in the number of GHB prescriptions for Medicaid and Medicare patients in 2019. Cost analysis identified a steady rise in the total cost of both GHB prescriptions and Medicaid program spending dedicated to GHB prescriptions prior to and during the pandemic years. This investigation along with future related research could allow for greater insight into how the COVID-19 pandemic has impacted GHB drug usage among Medicaid and Medicare Part B patients.

## Data Availability

All data produced in the present study are available upon reasonable request to the authors. All data produced are available online.

https://data.cms.gov/summary-statistics-on-use-and-payments/medicare-medicaid-spending-by-drug/medicaid-spending-by-drug

https://data.cms.gov/provider-summary-by-type-of-service/medicare-part-d-prescribers/medicare-part-d-prescribers-by-provider-and-drug

https://data.medicaid.gov/datasets?theme%5B0%5D=State%20Drug%20Utilization

## Acknowledgements

We would like to thank Elizabet Kuchinski, MPH and Jonique Depina MBS for their consistent support and guidance throughout this research project, Bhawana Chhetri for her contributions to data analyses, and Maria Tian for figure editing.

## Disclosures

Dr. Brian J. Piper was part of an osteoarthritis research team from 2019 to 2021, funded by Pfizer and Eli Lily. Software used in this report was provided by NIEHS (T32 ES007060-31A1). The other authors have no disclosures.

---

**Supplemental Figure 1:**
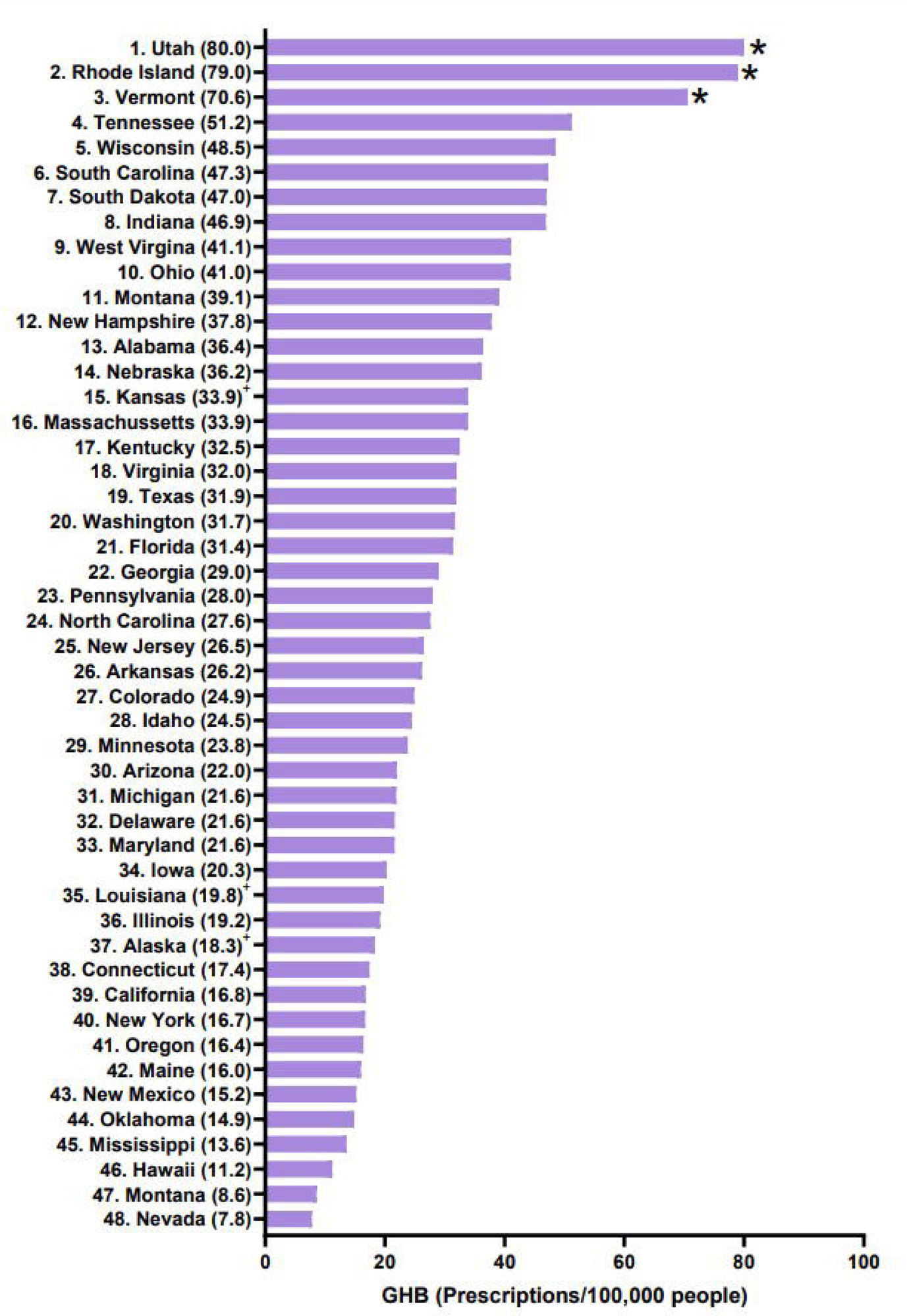
Scatterplot of GHB prescriptions per 100,000 patients for Medicaid and Medicare patients in 2019 (r(49) = -0.045, p = 0.7577).

**Supplemental Figure 2.**
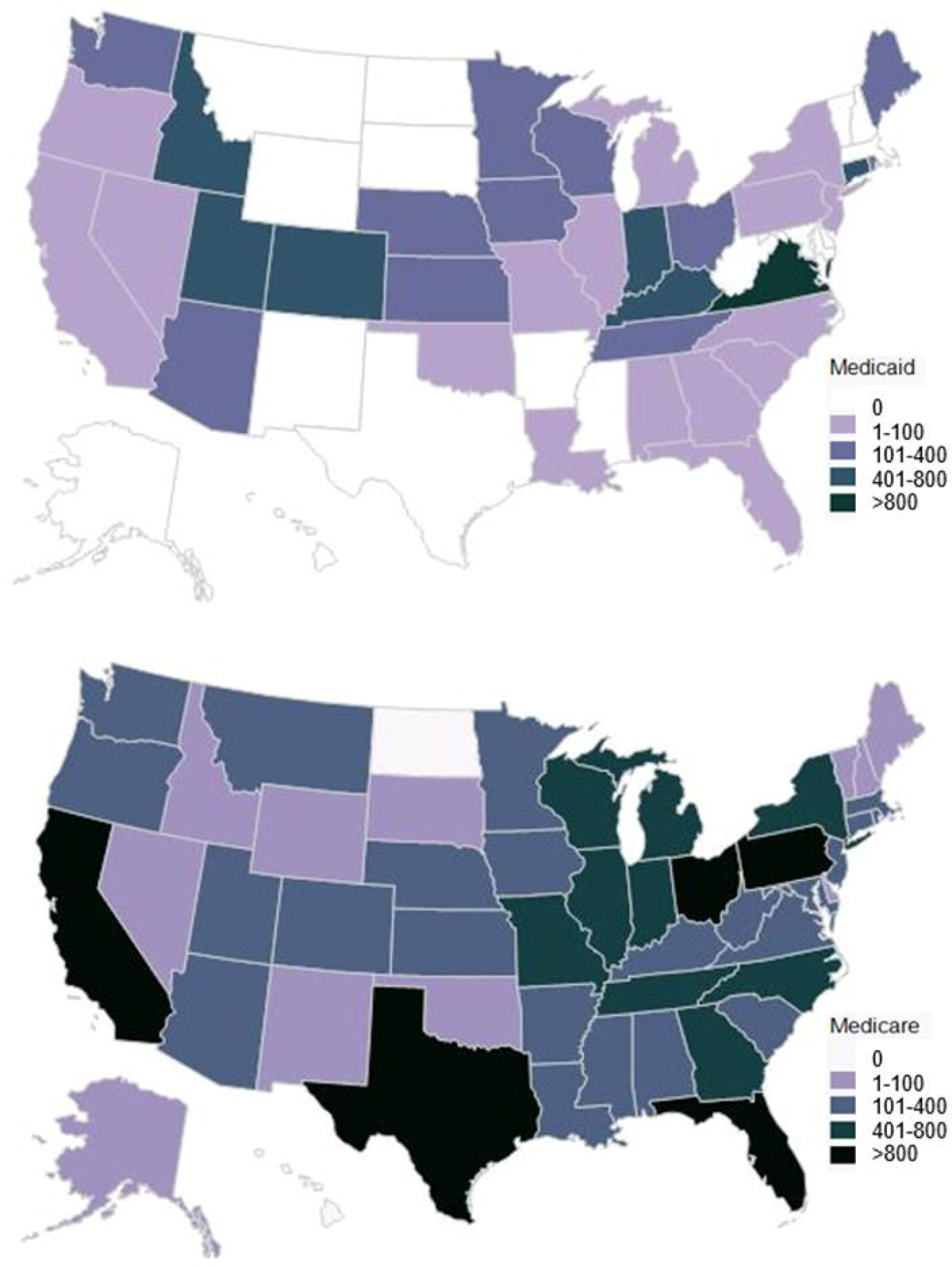
GHB prescription cost for Medicaid enrollees across a 5 year period (2017, 2018, 2019, 2020, 2021).

**Supplemental Figure 3.**
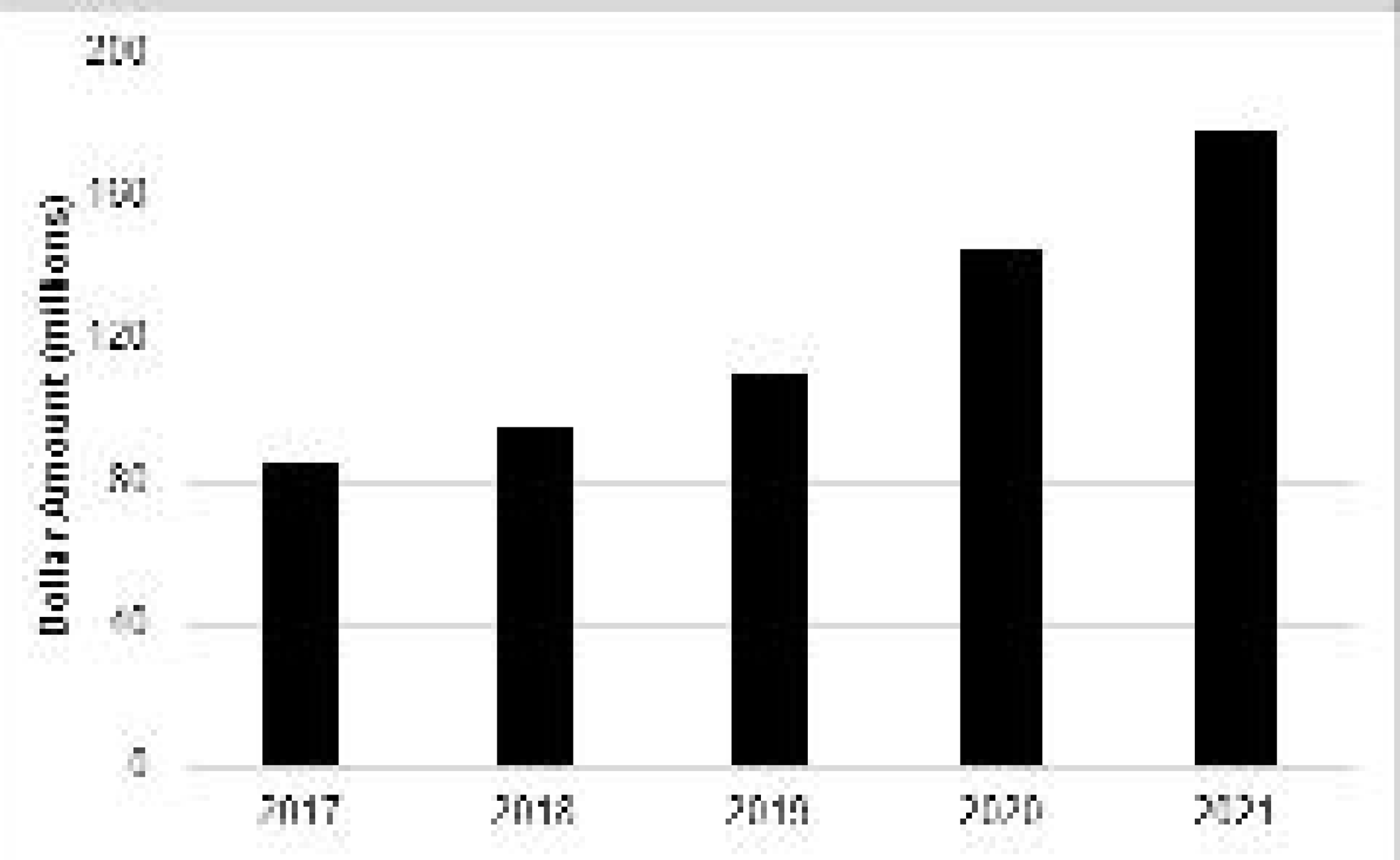
Heat maps of the US depicting the pronounced state-level disparities in gamma hydroxybutyrate (GHB) prescriptions in state per 100,000 patients enrolled in Medicaid (top) and Medicare (bottom) in 2019.

**Supplemental Figure 4.**
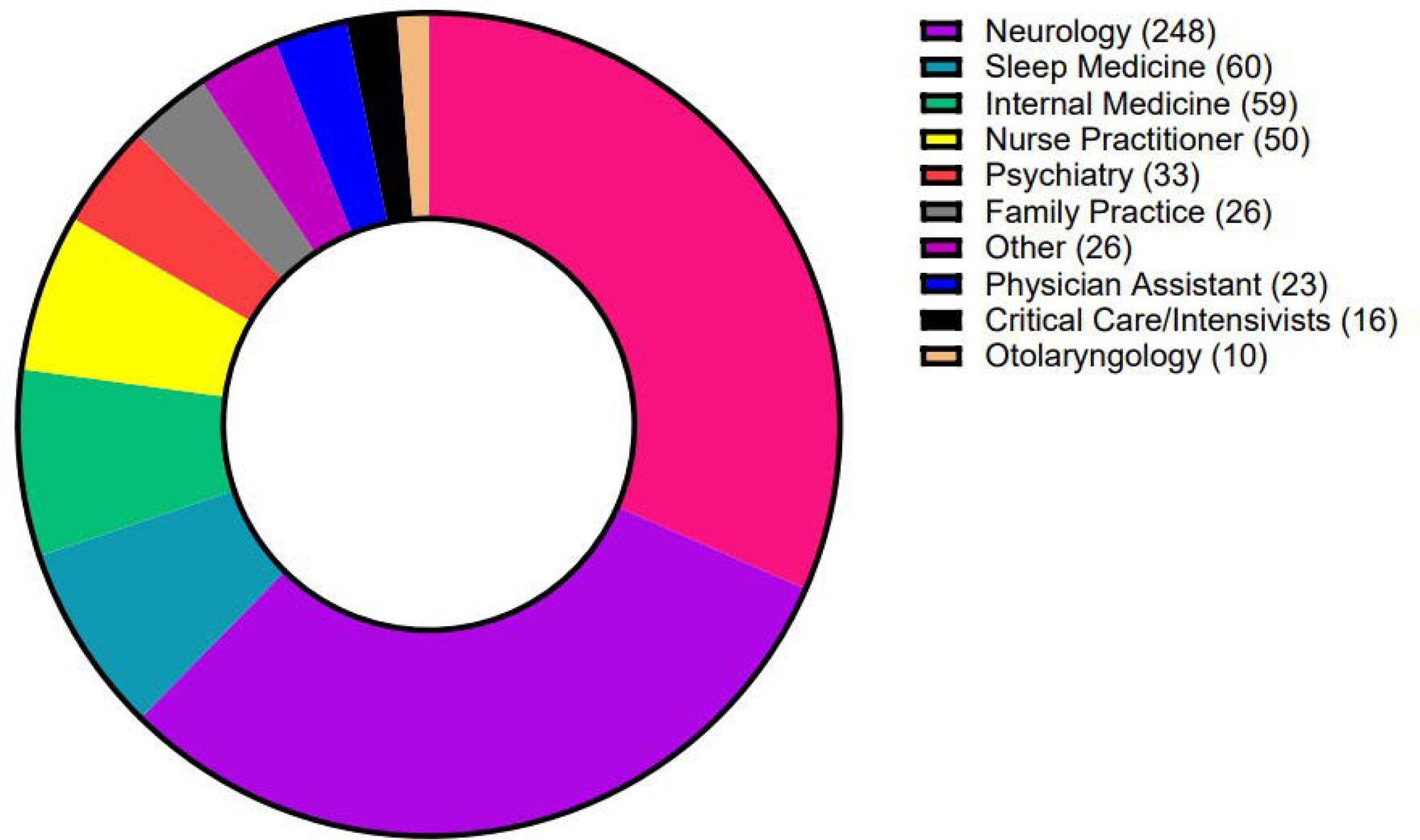
Heat maps of the US depicting the density of gamma hydroxybutyrate (GH)B prescriptions distributed across each state per 100,000 patients enrolled in Medicaid and Medicare in 2021.

**Supplemental Figure 5.**
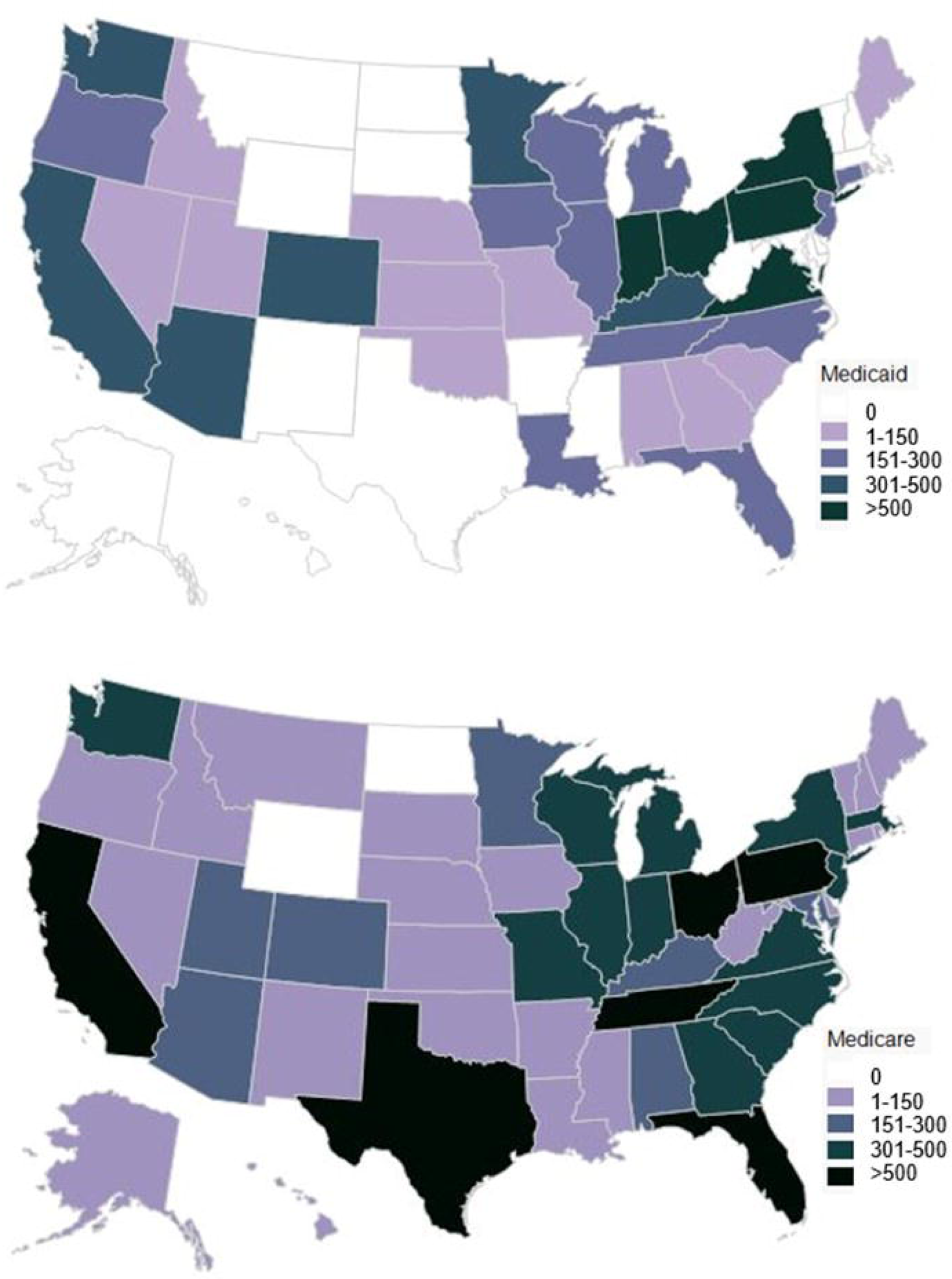
Heat maps of the US depicting the density of total number (i.e. uncorrected for population) of gamma-hydroxybutyrate (GHB) prescriptions per state for Medicaid and Medicare. The total number of GHB prescriptions prescribed in 2019.

**Supplemental Figure 6.**
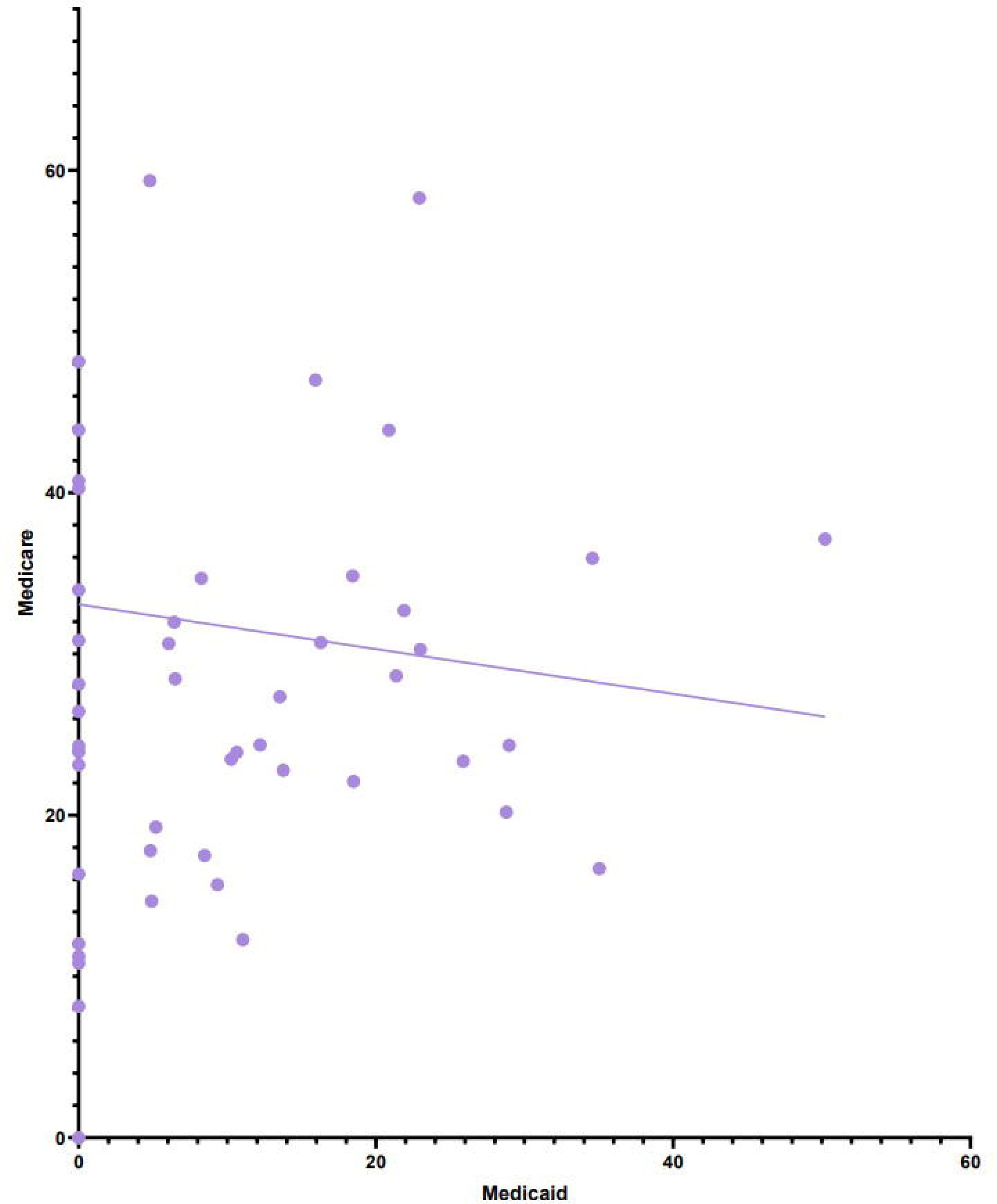
Heat maps of the US depicting the density of total (i.e. uncorrected for number of enrollees) number of gamma-hydroxybutyrate (GHB) prescriptions per state for Medicaid (top) and Medicare (bottom) in 2021.

**Supplemental Figure 7.**
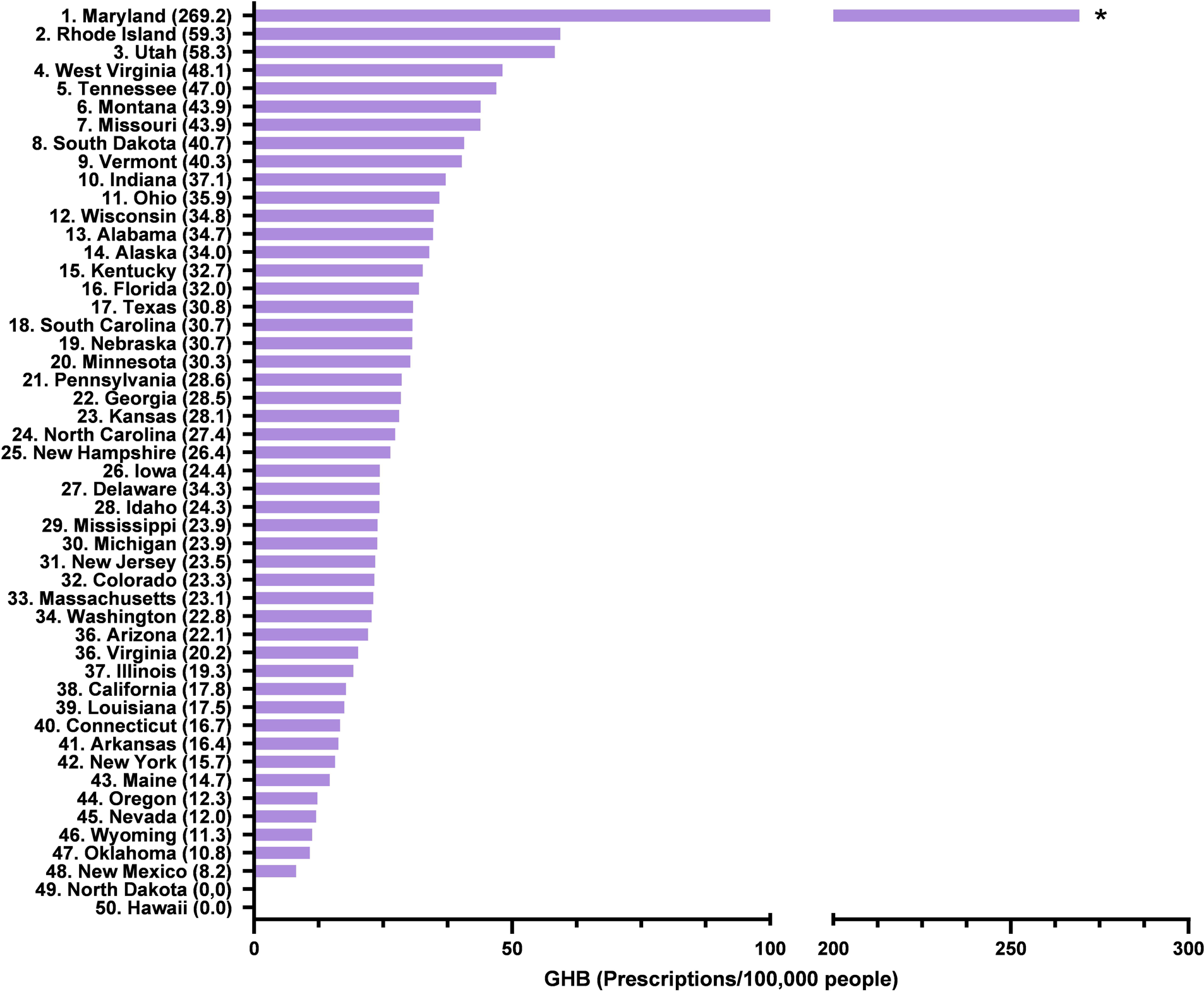
Gamma-hydroxybutyrate (GHB) prescribers for Medicare in 2019.

**Supplemental Table 1.**
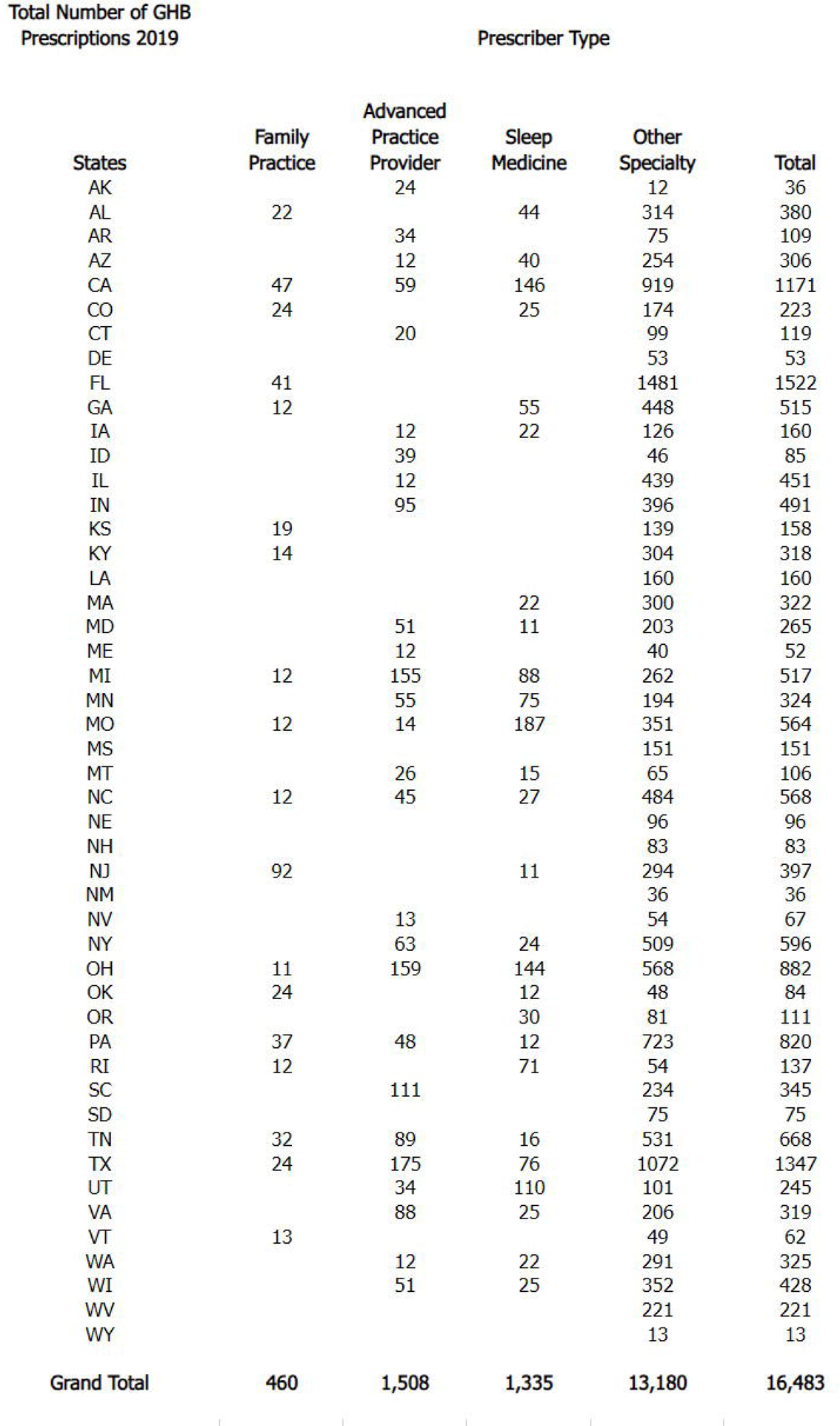
Data was extracted for all states from the State Drug Utilization tool on Medicaid.gov and from Data.CMS.gov. The total number of prescriptions per state from the 2019 year are listed on the right, and the total number of prescriptions from each database are listed on the bottom.

**Supplemental Table 2.** Data for all states was extracted from the Medicare Part D Provider and Drug dataset on Data.CMS.gov. The total number of prescriptions for Medicare enrollees were arranged based on the type of provider (family practice, advanced practice provider, other specialty, and sleep medicine). The bottom row depicts the total number of prescriptions given from each type of medical provider.

| Total Number of GHB Prescriptions | 2019 |  |
| --- | --- | --- |
|  | Medicaid | Medicare |
| AK | 0 | 36 |
| AL | 76 | 380 |
| AR | 0 | 109 |
| AZ | 316 | 306 |
| CA | 560 | 1,171 |
| CO | 330 | 223 |
| CT | 298 | 119 |
| DE | 67 | 53 |
| FL | 232 | 1,522 |
| GA | 118 | 515 |
| HI | 0 | 0 |
| IA | 83 | 160 |
| ID | 0 | 85 |
| IL | 146 | 451 |
| IN | 664 | 491 |
| KS | 0 | 158 |
| KY | 282 | 318 |
| LA | 125 | 160 |
| MA | 0 | 322 |
| MD | 0 | 265 |
| ME | 13 | 52 |
| MI | 247 | 517 |
| MN | 240 | 324 |
| MO | 177 | 564 |
| MS | 0 | 151 |
| MT | 0 | 106 |
| NC | 240 | 568 |
| ND | 0 | 0 |
| NE | 15 | 111 |
| NH | 0 | 83 |
| NJ | 175 | 397 |
| NM | 0 | 36 |
| NV | 0 | 67 |
| NY | 560 | 596 |
| OH | 902 | 882 |
| OK | 0 | 84 |
| OR | 110 | 111 |
| PA | 628 | 820 |
| RI | 14 | 137 |
| SC | 170 | 345 |
| SD | 0 | 75 |
| TN | 232 | 668 |
| TX | 0 | 1,347 |
| UT | 71 | 245 |
| VA | 407 | 319 |
| VT | 0 | 62 |
| WA | 238 | 325 |
| WI | 193 | 428 |
| WV | 0 | 221 |
| WY | 0 | 13 |
| Grand Total | 7,929 | 16,498 |

## Notes

### Funding Statement

This study did not receive any funding.

